# Visual Impairment and Risk of Dementia: the UK Biobank Study

**DOI:** 10.1101/2021.01.08.21249188

**Authors:** Zhuoting Zhu, Danli Shi, Huan Liao, Jason Ha, Xianwen Shang, Yu Huang, Xueli Zhang, Yu Jiang, Longyue Li, Honghua Yu, Wenyi Hu, Wei Wang, Xiaohong Yang, Mingguang He

**Author notes:** **Corresponding author** Mingguang He, MD PhD, Xiaohong Yang, MD PhD, Wei Wang, MD PhD.

## Abstract

**INTRODUCTION:** The association between visual impairment (VI) and the risk of dementia has been poorly understood. We sought to investigate the VI-dementia relationship in the UK Biobank Study.

**METHODS:** A total of 117,187 volunteers (aged 40-69 years) deemed free of dementia at baseline were included. Habitual distance visual acuity worse than 0.3 logMAR units in the better-seeing eye was used to define VI. The incident dementia was based on electronically linked hospital inpatient and death records.

**RESULTS:** During a median follow up of 5.96 years, the presence of VI was significantly associated with incident dementia (HR=1.78, 95% CI: 1.18-2.68, P=0.006). There was a clear trend between the severity of VI and the risk of dementia (P for trend=0.002).

**DISCUSSION:** Visually impaired individuals were more likely to develop incident dementia, with a progressively greater risk among those with worse visual acuity. Our findings highlight the value of regular vision screening and elimination of VI.

**HIGHLIGHTS:**

1. The association between VI and dementia has been poorly understood;
2. VI is associated with incident dementia in non-demented adults;
3. There is a clear trend between the severity of VI and the risk of dementia;
4. VI may be a marker of increased dementia risk.

**RESEARCH IN CONTEXT:** *SYSTEMATIC REVIEW:* We searched and reviewed the literature using traditional sources (e.g., PubMed and GoogleScholar). While the association between VI and cognitive function/decline are increasingly studies, investigation of the association between VI and the risk of dementia has been largely overlooked.

*INTERPRETATION:* We found that visually impaired individuals were more likely to develop incident dementia, with a progressively greater risk among those with worse visual acuity. Our findings imply that VI may be an important marker of dementia.

*FUTURE DIRECTIONS:* These findings call for more studies to investigate (a) the role of visual acuity changes on the risk of dementia; (b) the relationship between other components of visual function and incident dementia; (c) the relationship between eye diseases and incident dementia; and (d) the potential benefits of vision rehabilitation on dementia prevention.

## Introduction

The number of individuals with visual impairment (VI) and blindness is projected to more than double by 2050 as a result of shifting demographics and ageing population.^1,2^ In addition to vision loss, visually impaired individuals are more likely to suffer from mental disorders (e.g., depression),^3^ social isolation,^4^ unintentional injuries,^5^ functional disabilities,^6^ and poor survival.^7, 8^

Dementia is an increasing threat to global health, affecting nearly 46.8 million people worldwide, with this number expected triple by 2050.^9^ Despite this heavy burden, effective treatments for dementia are yet to be developed. Therefore, prevention, early detection and management are essential to address the enormous burden of dementia.

There is mounting evidence that suggests that VI and cognitive decline may be closely related.^10-22^ Nevertheless, the association between VI and risk of incident dementia has been poorly understood.^23-31^ Besides, the relatively small sample size, focus of late-onset dementia, self-reported visual function and bias of identifying dementia cases in previous studies may mask the true association between VI and incident dementia.

Therefore, we aimed to investigate the association of objectively determined VI and VI severity with incident dementia in a large-scale sample aged 40-69 years from the UK Biobank Study.

## Methods

### Study Sample

The UK Biobank is a large-scale prospective cohort study, enrolling more than 500,000 individuals (40-69 years) from across the UK, with baseline recruitment taking place during 2006-2010. Details of the rationale, design, and assessments used in the UK Biobank Study have been previously described elsewhere.^32^ Briefly, approximately 9.2 million people aged 40-69 years in the UK’s National Health Service (NHS) residing near one of 22 assessment centers were invited. A total of 502,505 individuals (response rate of 5.5%) agreed to participate and visited the assessment centers. During the baseline assessment, participants answered comprehensive questionnaires, underwent physical measures, and provided biological samples. They also gave consent to link to their health-related records. Ophthalmic assessments, including logMAR visual acuity (VA), autorefraction, intraocular pressure (IOP), keratometry, corneal biomechanics, and spectral-domain Optical Coherence Tomography (OCT) imaging, were introduced to the baseline assessment in 2009 for 6 assessment centers.

The UK Biobank Study’s ethical approval had been granted by the National Information Governance Board for Health and Social Care and the NHS North West Multicenter Research Ethics Committee. Written informed consent was obtained from all participants.

### Visual Acuity Testing

The detailed procedure of VA testing for the UK Biobank Study has been described elsewhere.^33^ Presenting distance VA was measured with the habitual correction (if any) at 4 m using the logarithm of the minimum angle of resolution (logMAR) chart (Precision Vision, LaSalle, Illinois, USA) on a computer screen. Presenting VA was scored as the total number of correctly read lines, converted to logMAR units. In the present analysis, VI was defined as the presenting VA worse than 0.3 logMAR units (Snellen 20/40) in the better-seeing eye. Based on the presenting VA in the better-seeing eye, the severity of VI was classified as mild (0.3 < logMAR ≤ 0.6; 20/40 < Snellen ≤ 20/80), moderate (0.6 < logMAR ≤ 0.7; 20/80 < Snellen ≤ 20/100) and severe (0.7 < logMAR; Snellen < 20/100).

### Ascertainment of Incident Dementia

Dementia cases in the UK Biobank Study were ascertained by combining data from participants’ medical history and record linkage to hospital admissions data and the national death register. Detailed information regarding the algorithms used to combine data from different sources to identify dementia can be found at the following link: https://biobank.ndph.ox.ac.uk/showcase/showcase/docs/alg_outcome_dementia.pdf. In the present analysis of incident dementia, we excluded all participants who already had a diagnosis of dementia in the hospital admissions data or self-reported dementia at the point of study entry. Follow-up time was calculated as the duration between the date of the first assessment and censored at the date of incident dementia, date of death, date of loss to follow-up, or 1^st^ March 2016, whichever occurred earliest.

### Covariates

Factors known to be associated with dementia were included as potential confounders in the present analysis. These variables included age at baseline assessment, sex, ethnicity (recorded as white and non-white), Townsend deprivation indices (an area-based proxy measure for socioeconomic status), education attainment (recorded as college or university degree, and others), family history of dementia (a marker of biological vulnerability), smoking status (recorded as current/previous and never), physical activity level (recorded as above moderate/vigorous/walking recommendation or not), and comorbidities (depression, diabetes, hypertension, and hyperlipidemia).

Self-reporting and/or the score on the Patient Health Questionnaire (PHQ, the first two items) of at least 3 were used to identify participants with depression.^34^ Hypertension was defined to include those participants who had self-reported, or doctor-diagnosed hypertension, were taking antihypertensive drugs, or had a systolic blood pressure of at least 130 mmHg or a diastolic blood pressure of at least 80 mmHg averaged over two measurements. Diabetes was defined to include those participants who had self-reported or doctor-diagnosed diabetes mellitus, were taking anti-hyperglycaemic medications or using insulin, or had a glycosylated hemoglobin level of ≥6.5%. Hyperlipidemia was defined to include participants with doctor-diagnosed hyperlipidemia, were taking lipid-lowering drugs, or had a total cholesterol level ≥6.21 mmol/L.

### Statistical Analysis

Descriptive statistics, including means and standard deviations (SDs), numbers and percentages, were used to report baseline characteristics of study participants. The unpaired t tests were used to compare means between two groups on continuous variables, and Pearson’s χ^2^ tests to compare distributions between two groups on categorical variables. The log-rank test was used to compare distributions of incident dementia between VI and non-VI groups. Hazard ratios (HRs) and 95% confidence intervals (CIs) were estimated based on the Cox proportional hazards regression models. Age- and sex-adjusted Cox proportional hazards regression models were used to identify covariates strongly associated with incident dementia. The covariates that were found to be associated with incident dementia in the age- and sex-adjusted models were then adjusted in the multivariable models.

We performed sensitivity analysis classifying VI using two different cutoffs: a presenting VA worse than 0.6 logMAR units (Snellen 20/80) and worse than 0.7 logMAR units (Snellen 20/100) in the better-seeing eye. To reduce the likelihood of accidental inclusion of prevalent cases, we also conducted sensitivity analysis after excluding dementia cases whose date of first recorded diagnosis occurred within one years from the date of baseline assessment.

The proportional hazards assumption was satisfied graphically. All tests were two-sided, and statistical significance was set at a P value of less than 0.05. All analyses were performed using Stata version 13 (StataCorp).

### Data availability

The UK Biobank is an open-access resource to researchers through registration of proposed research. (see ukbiobank.ac.uk/register-apply). Access to the UK Biobank data was granted after registration. The application ID was 62443.

## Results

### Study Sample

Of the 502,505 participants enrolled in the baseline UK Biobank Study between 2006 and 2010, VA was measured in 117,252 (23.3%) participants. Supplementary Table 1 demonstrates the baseline characteristics of participants with and without VA data. In summary, participants with VA data tended to be slightly older and living in a socioeconomically deprived area compared with those without VA data. There was a similar proportion of men and women, but compared to those without VA data, there was a slightly lower proportion of individuals who were current/former smokers, a slightly higher representation of non-white ethnicity, college or university degree, family history of dementia, above physical activity recommendation, and comorbidities than those without VA data. In the analysis of incident dementia, we excluded participants diagnosed with dementia prior to baseline assessment (n=52) and those who self-reported at the baseline nurse interview (n=13). A total of 117,187 participants were included in the final analysis.

Among the 117,187 participants who were not diagnosed with dementia at the baseline assessment, the mean (SD) age was 56.8 (8.11) years with 54.4% being female. Other baseline characteristics for the study population are shown in Table 1. A total of 4,018 (3.43%) participants had VI and 113,169 (96.6%) did not have VI. Table 1 provides the baseline characteristics for the VI and comparator groups. Participants with VI were more likely to be older, of non-white ethnicity, living in a socioeconomically deprived area and less educated than those without VI. Relative to the comparison group, a slightly lower proportion of visually impaired participants had a family history of dementia, but a higher proportion had depression, diabetes mellitus, and hypertension.

**Table 1.** Baseline Characteristics of the Study Participants Stratified by Visual Impairment Status.

| Baseline Characteristics | Total | Non-VI Group | VI Group | OR (95% CI)<sup>a</sup> |
| --- | --- | --- | --- | --- |
| N | 117,187 | 113,169 | 4,018 | - |
| Age, mean (SD), yrs | 56.8 (8.11) | 56.8 (8.12) | 58.8 (7.57) | **1.03 (1.03-1.04)** |
| Gender, No. (%) |  |  |  |  |
| Female | 63,745 (54.4) | 61,533 (54.4) | 2,212 (55.0) | 1 [Reference] |
| Male | 53,442 (45.6) | 51,636 (45.6) | 1,806 (45.0) | 0.96 (0.90-1.02) |
| Ethnicity, No. (%) |  |  |  |  |
| White | 104,249 (89.0) | 100,945 (89.2) | 3,304 (82.2) | 1 [Reference] |
| Others | 12,938 (11.0) | 12,224 (10.8) | 714 (17.8) | **2.08 (1.91-2.26)** |
| Townsend index, mean (SD) | -0.93 (3.01) | -0.96 (3.00) | -0.12 (3.34) | **1.10 (1.09-1.11)** |
| Education level, No. (%) |  |  |  |  |
| College or university degree | 40,557 (34.6) | 39,504 (34.9) | 1,053 (26.2) | 1 [Reference] |
| Others | 76,630 (65.4) | 73,665 (65.1) | 2,965 (73.8) | **1.42 (1.32-1.53)** |
| Smoking status, No. (%) |  |  |  |  |
| Never | 64,601 (55.5) | 62,410 (55.5) | 2,191 (55.4) | 1 [Reference] |
| Former/current | 51,795 (44.5) | 50,035 (44.5) | 1,760 (44.6) | 0.96 (0.90-1.02) |
| Physical activity, No. (%) |  |  |  |  |
| Not meeting recommendation | 16,867 (17.7) | 16,292 (17.7) | 575 (18.4) | 1 [Reference] |
| Meeting recommendation | 78,268 (82.3) | 75,711 (82.3) | 2,557 (81.6) | 0.93 (0.85-1.02) |
| Family history of dementia, No. (%) |  |  |  |  |
| No | 99,228 (84.7) | 95,760 (84.6) | 3,468 (86.3) | 1 [Reference] |
| Yes | 17,959 (15.3) | 17,409 (15.4) | 550 (13.7) | **0.79 (0.72-0.87)** |
| History of depression, No. (%) |  |  |  |  |
| No | 105,197 (89.8) | 101,692 (89.9) | 3,505 (87.2) | 1 [Reference] |
| Yes | 11,990 (10.2) | 11,477 (10.1) | 513 (12.8) | **1.38 (1.26-1.52)** |
| History of diabetes, No. (%) |  |  |  |  |
| No | 109,346 (93.3) | 105,706 (93.4) | 3,640 (90.6) | 1 [Reference] |
| Yes | 7,841 (6.69) | 7,463 (6.59) | 378 (9.41) | **1.37 (1.23-1.53)** |
| History of hypertension, No. (%) |  |  |  |  |
| No | 29,972 (25.6) | 29,141 (25.8) | 831 (20.7) | 1 [Reference] |
| Yes | 87,215 (74.4) | 84,028 (74.2) | 3,187 (79.3) | **1.17 (1.08-1.27)** |
| History of hyperlipidemia, No. (%) |  |  |  |  |
| No | 62,918 (53.7) | 60,857 (53.8) | 2,061 (51.3) | 1 [Reference] |
| Yes | 54,269 (46.3) | 52,312 (46.2) | 1,957 (48.7) | 0.96 (0.90-1.03) |

SD = standard deviation; VI = visual impairment; OR = odds ratio; CI = confidence interval.
<sup>a</sup> Logistic regression models adjusted for age and gender.

### Incident Dementia

The median (interquartile range, IQR) follow-up duration post baseline enrollment was 5.96 (5.77-6.23) years. A total of 438 participants (0.37%) were ultimately classified as having incident dementia. The baseline characteristics stratified by dementia status at follow-up are shown in the Table 2. Age- and gender-adjusted Cox proportional hazards regression models showed that ethnicity, education attainment, Townsend deprivation index, physical activity level, family history of dementia, comorbidities (depression and diabetes mellitus) at baseline were significant risk factors for the incident dementia.

**Table 2.** Baseline Characteristics Stratified by Dementia Status at Follow-up.

| Baseline Characteristics | Non-Dementia Group | Dementia Group | HR (95% CI)<sup>a</sup> |
| --- | --- | --- | --- |
| N | 116,749 | 438 | - |
| Age, mean (SD), yrs | 56.8 (8.10) | 64.0 (5.13) | **1.18 (1.16-1.21)** |
| Gender, No. (%) |  |  |  |
| Female | 63,550 (54.4) | 195 (44.5) | 1 [Reference] |
| Male | 53,199 (45.6) | 243 (55.5) | **1.39 (1.15-1.68)** |
| Ethnicity, No. (%) |  |  |  |
| White | 103,857 (89.0) | 392 (89.5) | 1 [Reference] |
| Others | 12,892 (11.0) | 46 (10.5) | **1.49 (1.10-2.02)** |
| Townsend index, mean (SD) | -0.93 (3.01) | -0.76 (3.17) | **1.06 (1.03-1.09)** |
| Education level, No. (%) |  |  |  |
| College or university degree | 40,471 (34.7) | 86 (19.6) | 1 [Reference] |
| Others | 76,278 (65.3) | 352 (80.4) | **1.73 (1.36-2.19)** |
| Smoking status, No. (%) |  |  |  |
| Never | 64,394 (55.5) | 207 (47.7) | 1 [Reference] |
| Former/current | 51,568 (44.5) | 227 (52.3) | 1.12 (0.93-1.36) |
| Physical activity, No. (%) |  |  |  |
| Not meeting recommendation | 16,788 (17.7) | 79 (24.7) | 1 [Reference] |
| Meeting recommendation | 78,027 (82.3) | 241 (75.3) | **0.58 (0.45-0.75)** |
| Family history of dementia, No. (%) |  |  |  |
| No | 98,897 (84.7) | 331 (75.6) | 1 [Reference] |
| Yes | 17,852 (15.3) | 107 (24.4) | **1.45 (1.16-1.80)** |
| History of depression, No. (%) |  |  |  |
| No | 104,841 (89.8) | 356 (81.3) | 1 [Reference] |
| Yes | 11,908 (10.2) | 82 (18.7) | **2.75 (2.16-3.50)** |
| History of diabetes, No. (%) |  |  |  |
| No | 108,988 (93.4) | 358 (81.7) | 1 [Reference] |
| Yes | 7,761 (6.65) | 80 (18.3) | **2.30 (1.80-2.94)** |
| History of hypertension, No. (%) |  |  |  |
| No | 29,909 (25.6) | 63 (14.4) | 1 [Reference] |
| Yes | 86,840 (74.4) | 375 (85.6) | 1.14 (0.87-1.50) |
| History of hyperlipidemia, No. (%) |  |  |  |
| No | 62,738 (53.7) | 180 (41.1) | 1 [Reference] |
| Yes | 54,011 (46.3) | 258 (58.9) | 1.08 (0.89-1.31) |

SD = standard deviation; HR = hazard ratio; CI = confidence interval.
<sup>a</sup> Cox proportional hazards regression models adjusted for age and gender.

Participants with VI at baseline were more likely to develop dementia at follow-up compared with those without VI (0.87% vs 0.36%, log-rank test, z = 28.5, P < 0.001). The age- and gender-adjusted Cox proportional hazard regression model found that VI was associated with a 2.01-fold higher risk of subsequent dementia (95% CI: 1.43-2.85, P < 0.001, Table 3). After further adjusting for potential confounders, the presence of VI was independently associated with a 78% higher risk of incident dementia (95% CI: 1.18-2.68, P = 0.006). There was a significant trend toward higher dementia risk across the VI severity groups (P for trend = 0.002), where the greatest risk for dementia was among those with severe VI (HR=3.53, 95% CI: 1.31-9.49, P = 0.013).

**Table 3.** Cox Proportional Hazards Models for Incident Dementia by Visual Impairment Status.

| VI Status | Age- and Gender-adjusted Model |  | Multivariable Model <sup>a</sup> |  |
| --- | --- | --- | --- | --- |
|  | HR (95% CI) | P value | HR (95% CI) | P value |
| VI status |  |  |  |  |
| None | 1 [Reference] | - | 1 [Reference] | - |
| VI | 2.01 (1.43-2.85) | <0.001 | 1.78 (1.18-2.68) | 0.006 |
| VI severity |  |  |  |  |
| None | 1 [Reference] | - | 1 [Reference] | - |
| Mild VI | 2.00 (1.38-2.91) | <0.001 | 1.66 (1.05-2.62) | 0.002 |
| Moderate VI | 0.91 (0.13-6.48) |  | 1.12 (0.16-8.01) |  |
| Severe VI | 3.05 (1.14-8.18) |  | 3.53 (1.31-9.49) |  |

VI = visual impairment; HR = hazard ratio; CI = confidence interval.
<sup>a</sup> Cox proportional hazards regression models adjusted for age, gender, race/ethnicity, education level, Townsend index, physical activity level, family history of dementia, history of diabetes mellitus, hypertension, and depression.

### Sensitivity Analysis

We observed similar findings of sensitivity analyses using different thresholds for classifying VI, where the risk of dementia was greatest among participants with a presenting VA worse than 0.7 logMAR units for VI (HR=3.42, 95% CI: 1.27-9.19, P=0.015), followed by a cutoff of 0.6 logMAR units for VI (HR=2.39, 95% CI: 0.99-5.80, P=0.052, Supplementary Table 2). Sensitivity analysis excluding participants with dementia cases firstly diagnosed within one year after the baseline assessment showed comparable results to the main analysis, again verifying the robustness of our findings (Supplementary Table 3).

## Discussion

In this large community-based population of 117,187 initially dementia-free individuals, we found that visually impaired participants were more likely to develop incident dementia, even after adjusting for potential confounders. We also found that the increased risk of dementia occurred in a graded fashion by VI severity. Our findings highlighted the value of regular vision screening and elimination of VI among middle-aged and elderly individuals.

Our findings have been corroborated by previous studies. The English Longitudinal Study of Ageing,^23^ the Gingko Evaluation of Memory Study,^24^ Health and Retirement Study cohort,^25^ and Women’s Health Initiative^26^ reported that the presence of VI was independently associated with a higher incidence of dementia during follow-up periods of 7 to 18 years. Interestingly, the Three-City study suggested that poor vision was an indicator of dementia risk at short (the first 2 years after inclusion) and middle-term (from 2 to 4 years after inclusion), but not at long-term (beyond 4 years after inclusion).^27^ Data from the Korean National Health Insurance Service^28^ and the cohort of the Hong Kong Elderly Health Centres^29^ showed that individuals with severe VI were at the greatest risk of incident dementia, thereby further supporting the association between VI and incident dementia. Nevertheless, other studies presented data refuting these conclusions: a case-control study in Germany, Michalowsky et al. found that VI was not significantly associated with the risk of dementia.^30^ Likewise, the association between poor visual acuity and the risk of developing dementia did not reach significance in the Health, Aging, and Body Composition Study.^31^ Discrepancy in study design, statistical methodology, demographic characteristics, definition of VI or poor vision, as well as the detection of dementia may partly explain the inconsistency among studies’ results.

Several mechanisms have been postulated to explain the association between VI and the risk of dementia, either in favor of VI being the manifestation of neurodegeneration, greater cognitive load and lower cognitive efficiency, functional disabilities, or shared pathogenesis between VI and dementia. Firstly, it has been suggested that VI may be one of the first manifestations of dementia.^35^ Secondly, visually impaired individuals may place more stress on neural resources to optimally perform visual tasks, thus increasing the cognitive loads.^36^ Alternatively, based on the visual deprivation hypothesis, reduced visual inputs and stimulation may compromise the cognitive efficiency.^37^ It remains possible that the association between VI and dementia may be due to the intermediate factors, such as depression, functional disabilities, social isolation, and decreased engagement in physical activity and cognitively stimulating activities.^38-40^ Last but not the least, the shared pathogenesis between the causes of VI and dementia may also explain this association. Beta-amyloid deposits and genetic risk factors for dementia have been implicated in patients with age-related macular degeneration, one of the leading causes of VI.^41^

From a health policy perspective, our findings highlight the value of regular vision screening and elimination of VI. Firstly, vision assessment is a widely available and low-cost test. In addition, with the advent of the smartphone-based eye care services,^42^ the accessibility and uptake of visual acuity testing have greatly improved, particularly in the disadvantaged areas. Given the strong evidence of the association between VI and incident dementia, visual acuity testing may be a valuable tool to identify individuals at higher risk of cognitive decline and dementia. Secondly, our findings imply the potential benefits of the vision training and VI elimination in dementia prevention. It has been indicated that vision-related training (e.g., the field of view training) and low vision rehabilitation (e.g., cataract surgery) may improve cognitive function.^43, 44^ Further studies are needed to investigate the benefits of vision training and vision rehabilitation in dementia prevention.

Despite the advantages of large sample size, long-term follow-up, complete adjustment for confounders, the age range in the analytical sample involving cases of early-onset dementia, and the use of routinely updated health-related records identifying incident dementia in the present analysis, several limitations should be borne in mind. First, we only explored the association between baseline visual acuity and the risk of dementia. Further studies are needed to investigate the effects of visual acuity changes and other components of visual function (e.g., contrast sensitivity) on the risk of dementia. Second, causes of VI were not available in the present analysis, thus preventing us from investigating associations between specific causes of VI and the risks of dementia. Third, due to the observational design, we could not draw the causal inferences. Nevertheless, we observed similar results in the sensitivity analysis, where dementia cases identified within one year after baseline assessment were excluded. Chances of reverse causality might be trivial. Last but not the least, although a broad array of potential confounders were accounted for in the statistical analysis, we could not completely exclude residual confounding.

In summary, we found that visually impaired individuals were more likely to develop incident dementia, with a progressively greater risk among those with worse visual acuity. Our findings highlight the value of regular vision screening and vision rehabilitation. Further research is warranted to confirm our findings and further investigate the effects of vision rehabilitation on dementia prevention.

## Data Availability

The UK Biobank is an open-access resource to researchers through registration of proposed research. (see ukbiobank.ac.uk/register-apply). The application ID for the present study was 62443.

## Abbreviations and Acronyms

VI: visual impairment
NHS: National Health Service
VA: visual acuity
IOP: intraocular pressure
OCT: Optical Coherence Tomography
PHQ: Patient Health Questionnaire
SD: standard deviations
HR: hazard ratios
CI: confidence intervals
IQR: interquartile range

## Author Contributions

Study concept and design: Zhu ZT, Shi DL, Liao H, He MG, Yang XH.

Acquisition, analysis, or interpretation: All authors.

Drafting of the manuscript: Zhu ZT, Shi DL, Liao H, Shang XW.

Critical revision of the manuscript for important intellectual content: Jason Ha, Wang W, He MG, Yang XH.

Statistical analysis: Zhu ZT, Shi DL, Shang XW.

Obtained funding: He MG, Yang XH.

Administrative, technical, or material support: Zhu ZT, Shang XW, Wang W, He MG, Yang XH.

Study supervision: He MG, Yang XH.

