## Supplementary Table 1 for "Visual Impairment and Risk of Dementia: the UK Biobank Study"

Supplementary Table 1. Baseline Characteristics of Participants With and Without Visual Acuity Data in the UK Biobank Study

| **Baseline Characteristics** | **No. of Participants without VA Data** | **No. of Participants with VA Data** | **P Value** |
|---|---|---|---|
| N | 385,252 | 117,252 | - |
| Age, mean (SD), yrs | 56.4 (8.09) | 56.8 (8.11) | **<0.001** |
| Gender, No. (%) | | | |
| Female | 209,613 (54.4) | 63,769 (54.4) | 0.890 |
| Male | 175,639 (45.6) | 53,483 (45.6) | |
| Ethnicity, No. (%) | | | |
| White | 368,385 (95.6) | 104,310 (89.0) | **<0.001** |
| Others | 16,868 (4.38) | 12,942 (11.0) | |
| Townsend index, mean (SD), yrs | -1.40 (3.11) | -0.93 (3.01) | **<0.001** |
| Education level, No. (%) | | | |
| College or university degree | 120,583 (31.3) | 40,580 (34.6) | **<0.001** |
| Others | 264,670 (68.7) | 76,672 (65.4) | |
| Smoking status, No. (%) | | | |
| Never | 208,893 (54.5) | 64,629 (55.5) | **<0.001** |
| Former/current | 174,202 (45.5) | 51,832 (44.5) | |
| Physical activity, No. (%) | | | |
| Not meeting recommendation | 249,368 (81.2) | 78,308 (82.3) | **<0.001** |
| Meeting recommendation | 57,718 (18.8) | 16,874 (17.7) | |
| Family history of dementia, No. (%) | | | |
| No | 344,802 (89.5) | 99,276 (84.67) | **<0.001** |
| Yes | 40,451 (10.5) | 17,976 (15.33) | |
| History of depression, No. (%) | | | |
| No | 347,689 (90.3) | 105,244 (89.8) | **<0.001** |
| Yes | 37,564 (9.75) | 12,008 (10.2) | |
| History of diabetes, No. (%) | | | |

| | | | |
|---|---|---|---|
| No | 362,834 (94.2) | 109,409 (93.3) | **<0.001** |
| Yes | 22,419 (5.82) | 7,843 (6.69) | |
| History of hypertension, No. (%) | | | |
| No | 109,205 (28.4) | 29,984 (25.6) | **<0.001** |
| Yes | 276,048 (71.6) | 87,268 (74.4) | |
| History of hyperlipidemia, No. (%) | | | |
| No | 209,724 (54.4) | 62,941 (53.7) | **<0.001** |
| Yes | 175,529 (45.6) | 54,311 (46.3) | |

SD = standard deviation
