## Supplementary Table 2 for "Visual Impairment and Risk of Dementia: the UK Biobank Study"

Supplementary Table 2. Cox Proportional Hazards Models for Incident Dementia by Visual Impairment Status Using Different Thresholds.

| | **Age- and Gender-adjusted Model** | | **Multivariable Model [a]** | |
|---|---|---|---|---|
| **VI Status** | **HR (95% CI)** | **P value** | **HR (95% CI)** | **P value** |
| VI at 0.6 logMAR | | | | |
| None | 1 [Reference] | - | 1 [Reference] | - |
| VI | 2.00 (0.83-4.84) | 0.122 | 2.39 (0.99-5.80) | 0.053 |
| VI at 0.7 logMAR | | | | |
| None | 1 [Reference] | - | 1 [Reference] | - |
| VI | 2.95 (1.10-7.89) | 0.031 | 3.42 (1.27-9.19) | 0.015 |

VI = visual impairment; HR = hazard ratio; CI = confidence interval.

[a] Cox proportional hazards regression models adjusted for age, gender, race/ethnicity, education level, Townsend index, physical activity level, family history of dementia, history of diabetes mellitus, hypertension, and depression.
