## Supplementary Table 3 for "Visual Impairment and Risk of Dementia: the UK Biobank Study"

Supplementary Table 3. Cox Proportional Hazards Models for Incident Dementia by Visual Impairment Status After Excluding Dementia Cases Within One Years After the Baseline Assessment.

| **VI Status** | **Age- and Gender-adjusted Model** | | **Multivariable Model [a]** | |
|---|---|---|---|---|
| | **HR (95% CI)** | **P value** | **HR (95% CI)** | **P value** |
| VI status | | | | |
| None | 1 [Reference] | - | 1 [Reference] | - |
| VI | 1.99 (1.40-2.83) | <0.001 | 1.80 (1.19-2.72) | 0.005 |
| VI severity | | | | |
| None | 1 [Reference] | - | 1 [Reference] | - |
| Mild VI | 1.97 (1.35-2.88) | <0.001 | 1.68 (1.07-2.65) | 0.002 |
| Moderate VI | 0.93 (0.13-6.59) | | 1.14 (0.16-8.12) | |
| Severe VI | 3.11 (1.16-8.32) | | 3.57 (1.33-9.59) | |

VI = visual impairment; HR = hazard ratio; CI = confidence interval.

[a] Cox proportional hazards regression models adjusted for age, gender, race/ethnicity, education level, Townsend index, physical activity level, family history of dementia, history of diabetes mellitus, hypertension, and depression.
